# Growth Differentiation Factor-15 as a candidate biomarker in gynecologic malignancies: A meta-analysis

**DOI:** 10.1101/2020.07.07.20148221

**Authors:** Dipayan Roy, Anupama Modi, Manoj Khokhar, Manu Goyal, Shailja Sharma, Purvi Purohit, Puneet Setia, Antonio Facciorusso, Praveen Sharma

## Abstract

Growth Differentiation Factor-15 (GDF-15), though emerged as a novel marker in gynecological cancers, is yet to be recognized in clinical diagnostics. Eligible studies were sorted from multiple online databases, namely PubMed, Cochrane, ClinicalTrials.gov, Google Scholar, Web of Science, Embase, Scopus, LILACS, OpenGrey. From six studies, histopathologically diagnosed cases without prior treatment, and with diagnostic accuracy data for GDF-15 in gynecological cancers, were included. Our meta-analysis shows that GDF-15 has pooled diagnostic odds ratio of 12.74 at 80.5% sensitivity and 74.1% specificity, and an AUC of 0.84. Hence, GDF-15 is a potential marker to differentiate gynecological malignancy from non-malignant tumors.

## 1. Introduction

Cancers of the ovary, fallopian tube, body of uterus, cervix, vagina, and vulva come under the broad heading of cancers of the female reproductive system. They are one of the major causes of cancer-related mortality, accounting for 13.1% of age-standardized cancer-related deaths in females (1). The difficulty of detecting cancer in its early stages is the primary factor for poor clinical outcomes (2). Biomarkers contribute to the management of these cancers by pre-operative differentiation between benign and malignant pelvic masses, progression of malignancy, monitoring response to treatment and recurrence, and most importantly, attempting to detect disease at an earlier stage (3). To date, there are no such biomarkers widely used in clinical settings of gynecological malignancies.

Serum cancer antigen 125 (CA125) has been in use as a biomarker for clinical diagnosis and monitoring of treatment response in epithelial ovarian carcinoma (EOC), but its sensitivity as an independent marker is suboptimal in early-stage post-menopausal women (3-5). Endometrial cancer incidence is on the rise worldwide. Early diagnosis can significantly improve its outcome as 5-year survival is >90% in early-stage disease. Existing markers (e.g., leptin, adiponectin, and prolactin) for detection or monitoring are subject to hormonal and metabolic alterations and not unique to cancer development (6). Also, cervical cancer is one of the leading causes of female cancer-related mortality. Although the conventional tumor marker, squamous cell carcinoma is a useful prognostic marker, its role in early diagnosis is limited (7).

Human Growth Differentiation Factor-15 (GDF-15), also known as Macrophage Inhibitory Cytokine (MIC-1), non-steroidal anti-inflammatory drug (NSAID) activated gene (NAG-1), placental bone morphogenetic protein (PLAB), placental transforming growth factor-beta (PTGFB), prostate derived factor (PDF), and PL-74 is a member of the Transforming Growth Factor-β (TGF-β) superfamily (8, 9). Under normal conditions, GDF-15 is only found in large amounts in the placenta (10). It is also a stress-responsive cytokine that is not only highly expressed in inflammatory conditions but has emerged as a potential marker in cancer diagnosis and progression (9, 11, 12). Recently, several studies have demonstrated it to be involved in different gynecological malignancies such as EOC (4, 13-16), endometrial carcinoma (17), uterine sarcoma (18), and cervical cancer (19), where it is seen to be increased in serum or upregulated in tissue in case of malignant tumors compared to non-malignant ones. Hence, its applicability as a biomarker that can differentiate a benign mass from a malignant one and aid in the early detection of gynecological malignancies would be a significant advancement. Recently, Maeno et al. (20) developed a novel, flow-through membrane immunoassay-based measurement system for GDF-15 for the screening of uterine sarcoma. This chemiluminescence assay-based method correlated well with the GDF-15 measurements from ELISA (18). Nevertheless, the inconsistency of the data across different studies prevents us from pinpointing its clinical relevance in these patients. We have compiled original articles citing the role of GDF-15 in cancers of the female reproductive tract and performed a systematic analysis to evaluate its importance as a diagnostic biomarker.

## 2. Materials and Methods

### 2.1 Data sources and eligibility criteria

We conducted this study following the Meta-analysis Of Observational Studies in Epidemiology (MOOSE) and Preferred Reporting Items for a Systematic Review and Meta-analysis of Diagnostic Test Accuracy Studies (PRISMA-DTA) guidelines (21, 22). Two independent reviewers performed a selective literature search on several databases (Pubmed, Cochrane, ClinicalTrials.gov, Google Scholar, Web of Science, Embase, Scopus, LILACS, Opengrey) during 16^th^-20^th^ March 2019. All relevant articles were screened as per titles and abstracts and subsequently reviewed for eligibility after they were combined and imported to Rayyan QCRI software (23). MeSH terms or keywords used in the search were ((“Growth Differentiation Factor-15” OR “GDF-15” OR “Macrophage Inhibitory Cytokine-1” OR “MIC-1” OR “NSAID-activated gene-1” OR “NAG-1” OR “placental transforming growth factor-beta” OR “PTGFB” OR “placental bone morphogenetic protein” OR “PLAB” OR “prostate derived factor” OR “PDF” OR “PL-74”) AND (“Ovarian cancer” OR “Ovarian carcinoma” OR “Endometrial cancer” OR “Endometrial carcinoma” OR “Uterine cancer” OR “Uterine Carcinoma” OR “Cervical cancer” OR “Cervical carcinoma”) AND (“diagnosis” OR “sensitivity” OR “specificity” OR “prognosis” OR “outcome”)). References of identified articles and related reviews were manually searched for articles that may have been left out. The entire search strategy was carried out again latest on 3^rd^ April 2021 for new articles which may have been left out of the analyses. No such study was found.

### 2.2 Study selection

The selection procedure is schematically depicted according to the PRISMA 2020 flow diagram (Figure 1) (24). Full-text studies were included as long as they met the following criteria: (1) included cases that were first identified without prior treatment; (2) proven diagnosis by pathology; (3) studies reported diagnostic feature of GDF-15 in cancers of the female reproductive tract; (4) sufficient data for describing or calculating Sensitivity, Specificity, and Area Under Curve (AUC); (5) studies approved by an Ethics Committee or Institutional Review Board, i.e. studies must have mentioned that written informed consent had been taken from all subjects before inclusion. Accordingly, exclusion criteria were-(1) studies in which patient received therapy, (2) studies with insufficient data, and also, failure to contact the authors, (3) studies with <10 cases, (4) duplicate publications, (5) non-clinical research, animal studies, reviews, conference abstracts, case reports, meta-analyses.

**Figure 1:**
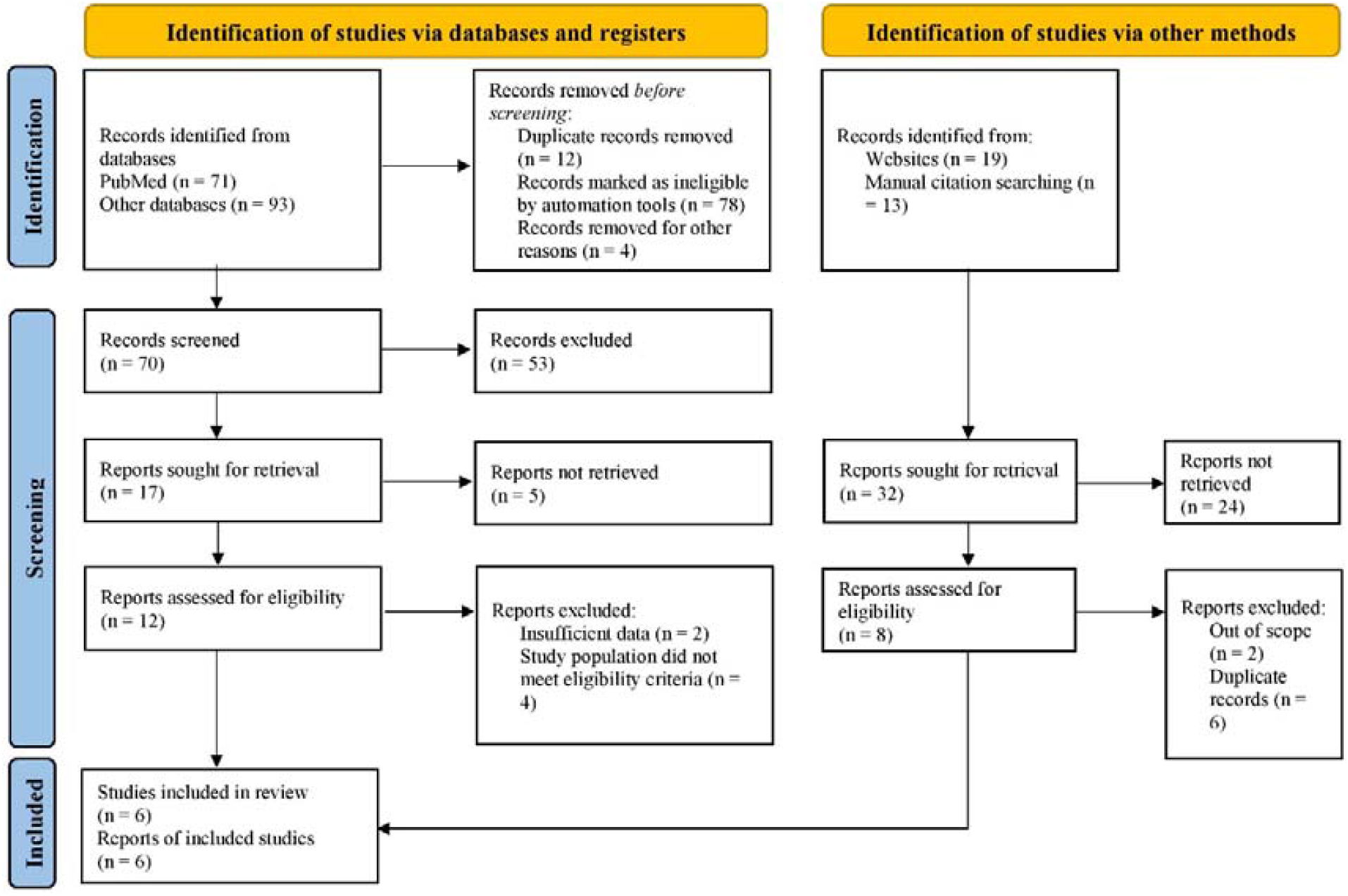
PRISMA 2020 flow diagram depicting the study selection procedure.

Registration detail: PROSPERO, CRD42019130097.

### 2.3 Data collection

Data were extracted from articles in the form of the lead author, year of publication, country of the study population, the number of patients, sample type, method of testing, sensitivity, specificity, cut-off value, and AUC by two independent reviewers. In the articles where sensitivity and specificity were not explicitly mentioned, they were extracted from the AUC curve using the Engauge Digitizer software (25) or manually calculated from other measures of diagnostic accuracy available in the articles (26) or both. Efforts were made to contact the authors of the original articles for the missing data but to no avail. In cases where a study had data for multiple comparisons, only one comparison was chosen in the final analysis considering the relevance to our research topic, heterogeneity issues and to avoid a unit-of-analysis error (27). All disagreements were resolved by discussion or consensus with a third reviewer.

### 2.4 Assessment of risk of bias

Quality assessment and risk of bias of the studies were evaluated by the revised Quality Assessment of Diagnostic Accuracy Studies-2 (QUADAS-2) criteria (28) according to the designated points viz. patient selection, index test, reference standard, and flow and timing. All relevant evidence were included in the final analysis (28).

### 2.5 Data analysis

Previously published guidelines and methods for conducting a meta-analysis of diagnostic test accuracy were consulted (29-31). Analyses were performed on Review Manager 5.3 (RevMan 5.3) (32), R programming platform with the aid of R packages **meta** and **mada** (Meta-Analysis of Diagnostic Accuracy) (33-37) and the MetaDTA online tool as described by Freeman et al. (38). The primary outcomes were obtained as pooled sensitivity, specificity, positive likelihood ratio (PLR), negative likelihood ratio (NLR), diagnostic odds ratio (DOR), and AUC with corresponding 95% confidence intervals (CI) by grouping the data from the studies. The bivariate model (39) for calculating summary measures in diagnostic accuracy studies was used for the analysis. Statistically significant heterogeneity among the studies was verified using Cochran’s Q and I^2^ statistics and a fixed or random-effects model was chosen accordingly. Funnel plot asymmetry for the examination of publication bias was not performed as the number of included studies was less than 10 (40).

For all analytical purposes, *p*<0.05 was considered statistically significant.

## 3. Results

### 3.1 Study selection

One hundred ninety-six (196), potentially relevant studies were retrieved in our search (Figure 1). They were scanned for titles, keywords, abstracts, and a total of one hundred eighty-four (184) studies were eliminated because they were either duplicates, out of scope or basic or animal model studies. The remaining 12 studies were read in detail, and finally, 6 of them, comprising 923 cases of gynecological cancers and 465 non-cancer controls, were considered eligible for diagnostic meta-analysis according to our inclusion criteria.

### 3.2 Study characteristics

The study characteristics are summarized in Table 1. In the six diagnostic studies finally included, the study participants involved Chinese (4, 15), Polish (13), Norwegian (14, 17, 18), and Belgian (18) patients, suffering from ovarian carcinoma (4, 13-15), endometrial carcinoma (17), and uterine sarcoma (18). The final diagnosis was all confirmed histologically and staged according to the International Federation of Gynecology and Obstetrics (FIGO) staging. The control group was non-cancer controls and comprised of healthy pre-and post-menopausal controls, benign ovarian tumors, and benign leiomyoma. Samples were either EDTA plasma (14, 17, 18), serum (4, 15), or peritoneal fluid (13). Enzyme-linked Immunosorbent Assay (ELISA) was primarily used to measure GDF-15, while one group each used immunoradiometric assay (IRMA) and multiplex immunoassay (13, 14). The diagnostic measures were sensitivity, specificity, and AUC. Only three out of the six studies provided a cut-off value of GDF-15, ranging from 519.6 pg/mL to 748 pg/mL (4, 14, 15).

**Table 1:** Study characteristics of all the articles included in the diagnostic meta-analysis.

| Study | Country and recruitment period | Design | Group1 (N1) | Group2 (N2) | Method | Cut-off (pg/mL) | TP | FN | FP | TN | Staging | Index test | Reference test |
| --- | --- | --- | --- | --- | --- | --- | --- | --- | --- | --- | --- | --- | --- |
| Staff 2010 (14) | Norway ; 2003-2009 | Prospective | Ovarian carcinoma (125) | Borderline and benign ovarian tumors (187) | EDTA Plasma; IRMA | 736 | 100 | 25 | 79 | 108 | 25/11/71/18 | GDF-15 | FIGO |
| Zhao 2018 (4) | China; 2009-2013 | Retrospective | Epithelial ovarian carcinoma (122) | Healthy control (120) | Serum; ELISA | 519.6 | 104 | 18 | 14 | 106 | I & II = 13; III & IV = 109 | GDF-15 | FIGO |
| Zhang 2016 (15) | China; 2010-2013 | Retrospective | Epithelial ovarian carcinoma (120) | Healthy control (40) | Serum; ELISA | 748 | 91 | 29 | 7 | 33 | I & II = 41; III & IV = 104 | GDF-15 | FIGO |
| Trovik 2014 (18) | Multicenter (Norway, Belgium); 2001-2010 | Prospective case control | Uterine sarcoma (19) | Benign leiomyoma (50) | EDTA Plasma; ELISA | - | 16 | 3 | 6 | 34 | 9/3/3/4 | GDF-15 | FIGO |
| Staff 2011 (17) | Norway ; 2003-2009 | Nested case control | Endometrial carcinoma (510) | Healthy control (40) | EDTA Plasma; ELISA | - | 319 | 182 | 18 | 32 | 366/57/63/24 | GDF-15 | FIGO |
| Chudecka-Glaz 2015 (13) | Poland; NR | Retrospective | Ovarian carcinoma (36) | Benign tumors (38) | Peritoneal fluid; Multiplex | - | 34 | 2 | 17 | 21 | I & II = 10; III & IV = 26 | GDF-15 | FIGO |

*Abbreviations:* CA125 = Cancer Antigen 125; ELISA = enzyme linked immunosorbent assay; FIGO = International Federation of Gynecology and Obstetrics; GDF-15 = Growth Differentiation Factor-15; IHC = Immunohistochemistry; IRMA = Immunoradiometric assay; NR = not reported

### 3.3 Risk of bias of included studies

The studies were assessed for quality using the QUADAS-2 list (Table 3). According to the QUADAS-2 assessment, five out of six studies were at risk of bias, whereas only one study showed concern regarding applicability. All studies were included for further statistical analysis.

### 3.4 Synthesis of results

The Higgins’ I^2^ for the diagnostic odds ratio of all studies was 86.5% (95% CI 72.8%-93.3%, *p*<0.01), indicating considerable heterogeneity in the pooled data. Values ranging from 60.9% to 90.6% were also detected in other subgroups in the diagnostic data. Hence, we resorted to using the random-effects model for the studies.

The measures of diagnostic accuracy for the included studies are depicted in Table 2 and Figure 2. The overall pooled sensitivity, specificity, diagnostic odds ratio (DOR) and area under the curve (AUC) for GDF-15 used to distinguish gynecological cancers from non-cancerous tumors, were 0.805 (95% CI: 0.711-0.873), 0.741 (95% CI: 0.611-9.839), 12.738 (95% CI: 5.034-32.231), and 0.835 respectively, corresponding to a positive likelihood ratio (PLR) of 3.103 (95% CI: 1.671-4.535) and a negative likelihood ratio (NLR) of 0.264 (95% CI: 0.146-0.383). The summary ROC curve is shown in Figure 3. These results suggest that GDF-15 levels can be used as a useful alternative biomarker in diagnosing cancers of the female reproductive tract compared to non-cancerous tumors.

**Table 2:** Measures of diagnostic accuracy in the selected studies.

| Study | Sensitivity<br>(95% CI) | Specificity<br>(95% CI) | DOR (95% CI) | AUC |
| --- | --- | --- | --- | --- |
| Staff 2010 (14) | 0.800 (0.719;<br>0.866) | 0.578 (0.503;<br>0.649) | 5.47 (3.23; 9.25) | 0.790 |
| Zhao 2018 (4) | 0.853 (0.777;<br>0.910) | 0.883 (0.812;<br>0.935) | 43.75 (20.69; 92.52) | 0.913 |
| Zhang 2016 (15) | 0.758 (0.672;<br>0.832) | 0.825 (0.672;<br>0.927) | 14.79 (5.92; 36.99) | 0.894 |
| Trovik 2014 (18) | 0.842 (0.604;<br>0.966) | 0.850 (0.702;<br>0.943) | 30.22 (6.69; 136.52) | 0.710 |
| Staff 2011 (17) | 0.637 (0.593;<br>0.679) | 0.640 (0.492;<br>0.771) | 3.12 (1.70; 5.71) | 0.860 |
| Chudecka-Glaz<br>2015 (13) | 0.944 (0.813;<br>0.993) | 0.553 (0.383;<br>0.714) | 21.00 (4.40; 100.22) | 0.752 |
| Pooled data | 0.805 (0.711;<br>0.873) | 0.741 (0.611;<br>0.839) | 12.74 (5.03; 32.23) | 0.835 |

*Abbreviations:* 95% CI = 95% Confidence Interval; AUC = Area under the curve; DOR =
Diagnostic odds ratio

**Table 3:** Evaluation of the risk of bias and applicability concerns of the included diagnostic studies using QUADAS-2

| Study | Risk of Bias |  |  |  | Applicability Concerns |  |  |
| --- | --- | --- | --- | --- | --- | --- | --- |
|  | Patient Selection | Index Test | Reference Standard | Flow and Timing | Patient Selection | Index Test | Reference Standard |
| Staff 2010 (14) | Low | Low | Low | Low | Low | Low | Low |
| Zhao 2018 (4) | High | Low | Low | Low | Low | Low | Low |
| Zhang 2016 (15) | High | Unclear | Low | Low | Low | Low | Low |
| Trovik 2014 (18) | Unclear | Low | Low | Low | Low | Low | Low |
| Staff 2011 (17) | High | Low | Low | Low | Low | Unclear | Low |
| Chudecka-Glaz 2015 (13) | Low | Unclear | Low | Low | Low | Low | Low |

**Figure 2:**
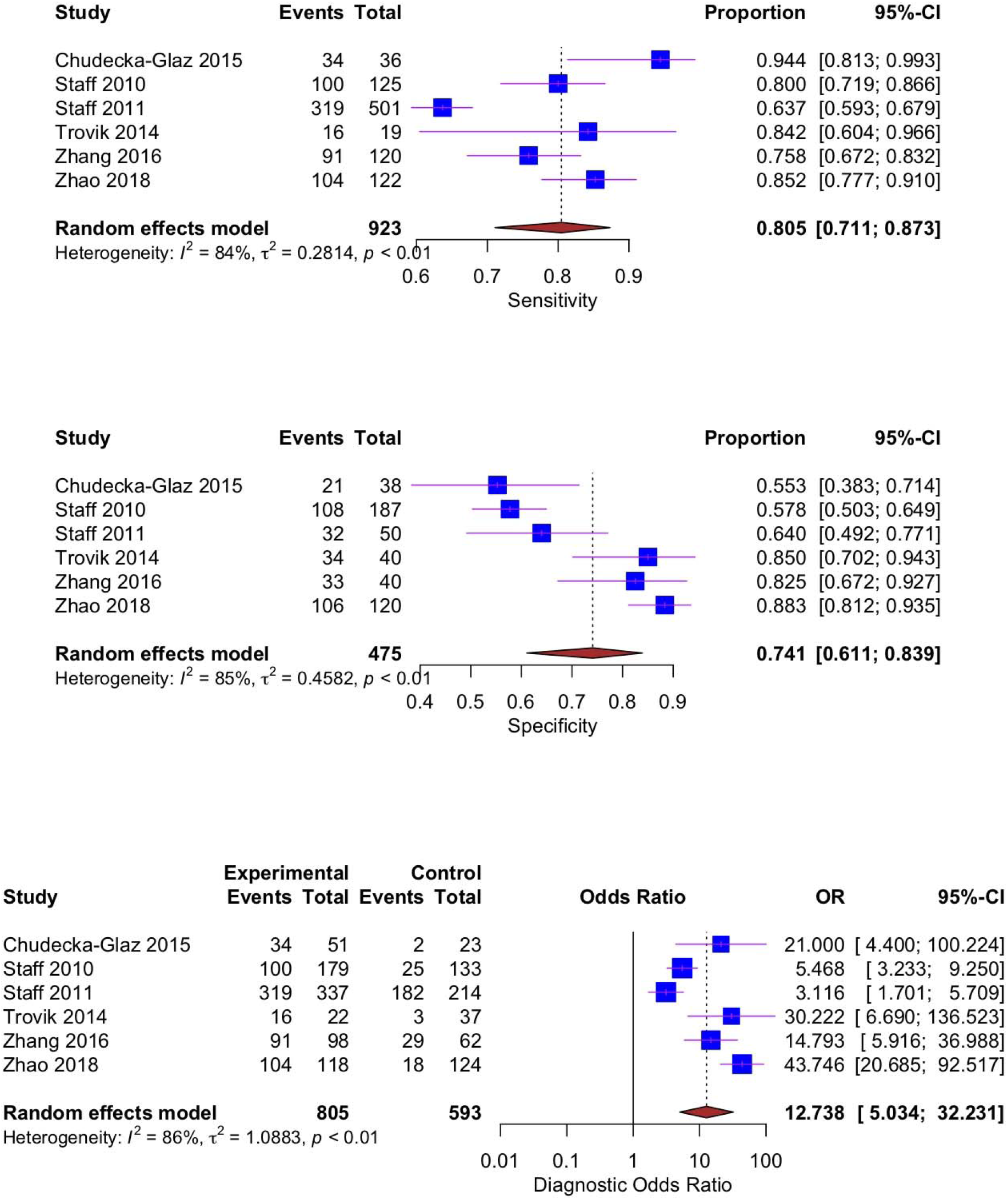
Forest plots of GDF-15 for the (a) sensitivity, (b) specificity, and (c) diagnostic odds ratio of the pooled data from the included studies

**Figure 3:**
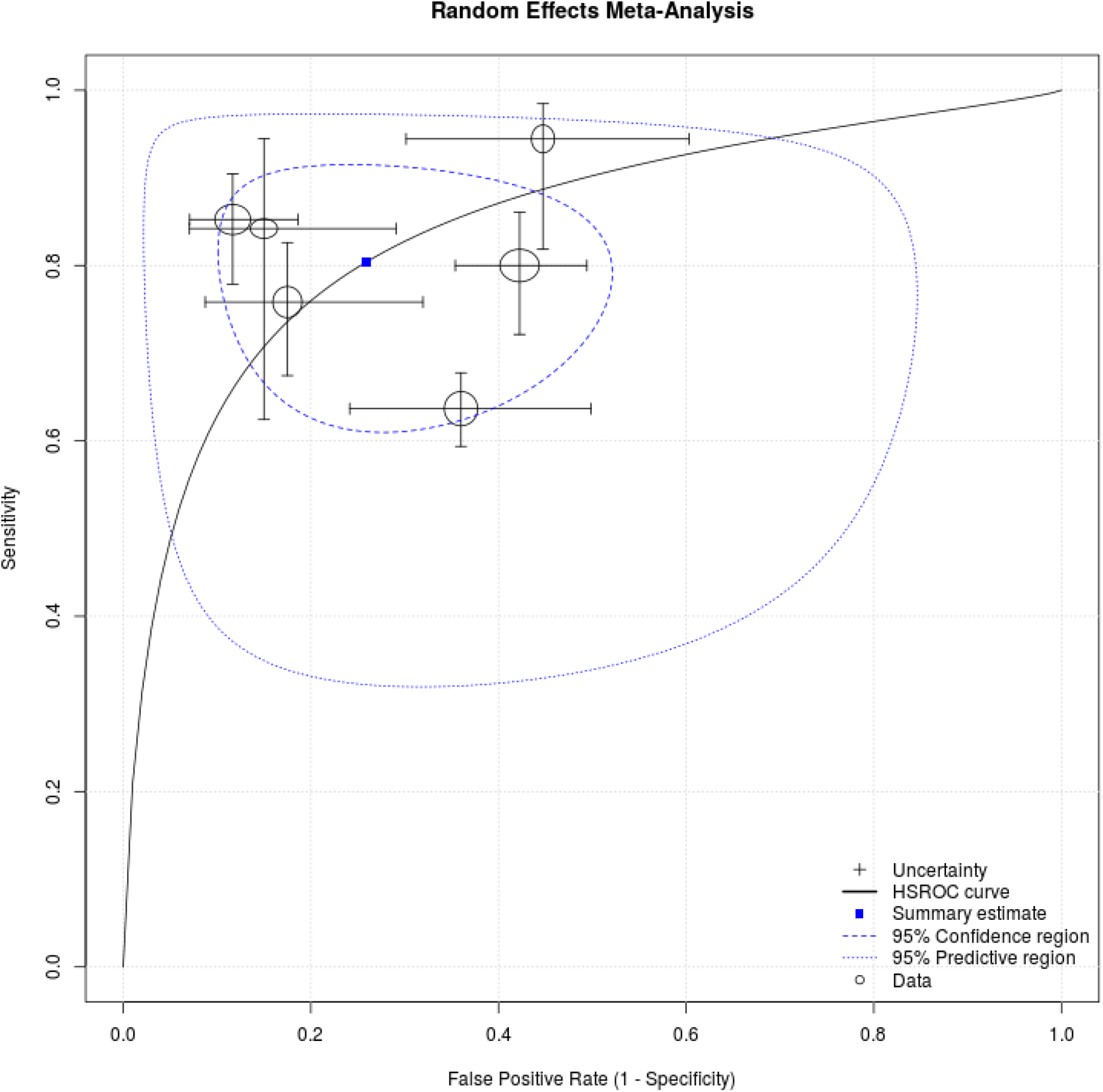
ROC plot displaying extrapolated SROC curve, the summary estimate for sensitivity and specificity, and percentage study weights

## 4. Discussion

Our study has identified six original articles which evaluated the role of GDF-15 as a candidate biomarker in diagnosing malignancies of the female reproductive tract from non-malignant tumors. We performed a meta-analysis and found it to be a potential robust biomarker with a pooled sensitivity and specificity of 80.5% and 74.1%, respectively. One study (14) had a ‘low risk’ of bias and applicability across all domains in the QUADAS-2 assessment. Five other studies had either ‘unclear’ or ‘high’ risk of bias in one of the four domains, among which Staff et al. (17) also had one ‘unclear’ applicability concern. We also found significant heterogeneity across the studies.

Malignancies of the female reproductive tract are one of the major causes of cancer-related mortality in females, mainly because these cancers are diagnosed at later stages (1, 2). Pre-operative detection of malignant lesions is also crucial for optimal management. Furthermore, the disease-free survival (DFS) of EOC patients is often poor even after extensive resection (41). Several systematic reviews in the past have elaborated the role of existing biomarkers or combinations thereof in ovarian or endometrial cancers alone (42-45). CA125 as a diagnostic biomarker to differentiate malignant and borderline ovarian tumors from benign lesions showed a promising sensitivity and specificity of 80% and 75%, respectively (45), which is comparable to our pooled estimates. Nevertheless, to our knowledge, a marker potent enough to discriminate malignant gynecological tumors in early stages from non-malignant ones without compromising on either sensitivity or specificity is not yet described. Several groups have proposed GDF-15 as a biomarker in prostatic, colorectal, hepatocellular, and pancreatic carcinomas as well as other malignancies and non-malignant inflammatory conditions (46, 47). Therefore, it is clear from the existing evidence that GDF-15 may not be specific for any single cancer type. However, GDF-15 is substantially increased or upregulated across all gynecological malignancies. Thus, it can be used as a complementary diagnostic biomarker in addition to existing diagnosing strategies, wherein it may be a useful biomarker in the diagnosis of early-stage EOC from benign ovarian tumors and other non-cancerous tumors of the female reproductive tract in such scenarios. In our study, the pooled sensitivity was comparably close with the sensitivity values of GDF-15 across all articles which were taken for analysis. While two articles on ovarian carcinoma showed the specificity of GDF-15 to be below 60% (13, 14), the pooled specificity came out to be 74.1%, possibly because of sizeable inter-study variation.

There are certain limitations to our study. There were low sample sizes for individual cancer types. Most of the studies were at risk of bias, primarily due to a case-control design, which is known to exaggerate diagnostic accuracy (48, 49). So far, the studies have been confined only to Chinese and European populations. Thus, a small sample with such a highly selective population will require further validation studies to establish its applicability in diagnosis in a clinical setting. Studies on a large scale need to be carried out in other populations. In our review, we could not include studies on all the cancers of the female reproductive system, namely cervical and vulval cancers, as data were either not present or did not fit our inclusion criteria. Furthermore, studies on in-vitro models and animal models were also left out. Patients with treatment history had to be excluded as anti-cancer therapy may alter the expression levels of GDF-15.

The applicability of GDF-15 can be verified with future prospective studies focusing on larger sample size and patient ethnicities. Finally, before implementing it into clinical practice, cut-off values for GDF-15 must be determined and internationally validated. Here, the cut-off value to differentiate between malignant and non-malignant tumors or healthy controls is mentioned in only three of the six studies included. Large scale prospective cohorts, then, are necessary to validate a uniform cut-off. To summarize, our analysis suggests that GDF-15 may be a useful candidate marker to differentiate malignant from non-malignant tumors of the female reproductive system.

## Data Availability

Not applicable.

## Acknowledgement

None.

